# Pregnancy during COVID-19: social contact patterns and vaccine coverage of pregnant women from CoMix in 19 European countries

**DOI:** 10.1101/2022.06.01.22275775

**Authors:** Kerry LM Wong, Amy Gimma, Enny S Paixao, CoMix Europe Working Group, Christel Faes, Philippe Beutels, Niel Hens, Christopher I Jarvis, W. John Edmunds

**Affiliations:** Department of Infectious Disease Epidemiology, London School of Hygiene and Tropical Medicine, Keppel Street, London WC1E 7HT, UK; Interuniversity Institute of Biostatistics and statistical Bioinformatics, Data Science Institute, Hasselt University, Hasselt, Belgium; Centre for Health Economic Research and Modelling Infectious Diseases, University of Antwerp, Antwerp, Belgium

## Abstract

**Background:** Evidence and advice for pregnant women evolved during the COVID-19 pandemic. We studied social contact behaviour and vaccine uptake in pregnant women between March 2020 and September 2021 in 19 European countries.

**Methods:** In each country, repeated online survey data were collected from a panel of nationally-representative participants. We calculated the mean adjusted contacts reported with an individual-level generalized additive mixed model, modelled using the negative binomial distribution and a log link function. Mean proportion of people in isolation or quarantine, and vaccination coverage by pregnancy status and gender were calculated using a clustered bootstrap.

**Findings:** We recorded 4,129 observations from 1,041 pregnant women, and 115,359 observations from 29,860 non-pregnant individuals aged 18-49. Pregnant women made slightly fewer contacts (3.6, 95%CI=3.5-3.7) than non-pregnant women (4.0, 95%CI=3.9-4.0), driven by fewer work contacts but marginally more contacts in non-essential social settings. Approximately 15-20% pregnant and 5% of non-pregnant individuals reported to be in isolation and quarantine for large parts of the study period.

COVID-19 vaccine coverage was higher in pregnant women than in non-pregnant women between January and April 2021. Since May 2021, vaccination in non-pregnant women began to increase and surpassed that in pregnant women.

**Interpretation:** Social contacts and vaccine uptake protect pregnant women and their newborn babies. Recognition of maternal social support need, and efforts to promote the safety and effectiveness of the COVID-19 vaccines during pregnancy are high priorities in this vulnerable group.

## Introduction

The physiological changes and relative immunodeficiency during pregnancy increase vulnerability to outbreaks of emerging infectious diseases for both the mother and foetus. Although being pregnant is not an additional risk for getting COVID-19, research findings show that, if infected, pregnant women are five times more likely to be hospitalized compared to non-pregnant women of a similar age^1^. A review of 192 studies showed that compared to pregnant women without COVID-19, those with the disease had increased odds of intensive care unit (ICU) admission (odds ratios (OR) of 18.6 (95%CI=7.5-45.8) and maternal death (OR=7.5, 95%CI=1.1-7.5). The odds of preterm birth (OR=1.5, 95%CI=1.1-1.9) and admission to neonatal ICU (OR=4.9, 95%CI=1.0-12.9) were also higher in babies born to mothers with COVID-19 versus those without^1^.

The World Health Organization urges for pregnant women and those around them to take precautions to protect themselves against COVID-19. In Europe, the National Health Service in the United Kingdom (UK) and European Centre for Disease Prevention and Control have advised pregnant women to avoid crowded and confined indoor places, limit the number of people they meet, and ask others to take a COVID-19 test before meeting up^2,3^. These guidelines, alongside other social distancing and non-pharmaceutical interventions (NPIs) implemented by governments for the public – e.g., restrictions on events and lockdown/”stay-at-home” orders – can reshape social behaviour and reduce individuals’ opportunities for social support and connection, a key buffer for risk of general and psychosocial wellbeing especially for expectant mothers. As the pandemic worsened in early spring 2020, pregnant women reported increased levels of stress, fear and uncertainty^4^. However, evidence on how pregnant women alter their contact patterns in response to covid-related recommendations and restrictions has remained scarce.

There has also been particular concern surrounding the development and authorisation of the COVID-19 vaccines that are safe to use in pregnancy. Across countries, guidelines with respect to recommending or withholding COVID-19 vaccines for pregnant women range from permissive (e.g., in the UK^5^) to more restrictive (e.g., Slovak Republic^6^). The variability in policy positions is partially due to limited evidence, as pregnant women were excluded from most trials conducted prior to authorization or listing, though efforts are underway to improve the evidence base.

In this analysis, we use primary data collected over the course of the COVID-19 pandemic in the European region to investigate the impact of the pandemic on the health of pregnant women in 19 countries. We describe the impact of the pandemic, focusing on social contact and social isolation, risk perception, and vaccination coverage over time and across various levels of government restrictions.

## Methods

### Ethics statement

Participation in this opt-in study was voluntary, and all analyses were carried out on anonymised data. In the UK, the study was approved by the ethics committee of the London School of Hygiene and Tropical Medicine (Reference number: 21795). In Belgium, the study was approved by Ethical Comimtte of the University Hospital Antwerp. In The Netherlands, waiver was obtained from Medische Ethische Toetsingscommissie Utrecht. In Austria, the study was waived by Ethikkommission der Stadt Wien. In Denmark, waiver was obtained from The Central Denmark Region Committees on Health Research Ethics. In France, waiver was obtained from Direction centrale du renseignement interieur Centre Hospitalier Universitaire de Poitiers. In Poland, the study was approved by Komisja Bioetyczna, Narodowego Instytutu Zdrowia Publicznego, Panstwowego Zaldadu Higieny, Warszawa. In Portugal, the study was approved by Comissao de Etica para a Saude do Instituto Nacional de Saude Doutor Ricardo Jorge. In Spain, the study was approved by Comite dEtica de la Investigacion del Hospital Universitari Germans Trias i Pujol. In Italy, the study was approved by Comitato di Bioetica d’ Ateneo, Universita degli studi di Torino. In Finland, waiver was obtained from Institutional Review Board of the Finnish Institute for Health and Welfare. In Switzerland, the study was approved by Gesundheits-, Sozial-und Integrationsdirektion Kantonale Ethikkommission fur die Forschung. In Lithuania, waiver was obtained from the Lithuanian Bioethics Committee. In Greece, the study was approved by Research Ethics Committee and University of West Attica. In Slovenia, the study was approved by Komisija Republike Slovenije za medicinsko etiko. In Croatia, the study was approved by Etiko povjerenstvo Hrvatskog zavoda za javno zdravstvo. In Estonia, the study was approved by Research Ethics Committee of the National Institute for Health Development. In Hungary, the study was approved by Health Science Council - Scientific and Research Ethics Committee Budapest. In Slovakia, the study was approved by Eticka komisia Univerzita Komenskeho v Bratislave Jesseniova lekarska fakulta v Martine. Further details are available from https://github.com/wongkerry/comix_preg/blob/main/ethics_committees.xlsx.

### Study design and setting

CoMix is an online longitudinal, multi-country social contact survey that follows individuals in 19 European countries over the course of the COVID-19 pandemic. The survey asks people aged 18 or above about their awareness, perceptions, social contacts, and health condition over the course of the COVID-19 pandemic. Study participants are invited to the survey and asked to respond every one to two week(s) apart. In each country, a nationally representative sample was recruited by the market research company Ipsos-MORI or a local vendor using quota sampling based on age, gender, and geographic region, and when possible, socioeconomic status to reflect the distribution within the population. Recruitment was conducted through web advertising and email campaigns.

The design of the CoMix survey is based on the POLYMOD contact survey^8^ with additional questions about work attendance, household composition, self-isolation and -quarantine due to COVID-19, and COVID-19 vaccination (since December 2020), among others. Details of the CoMix study including the protocol, further methodological details and survey instrument for the UK panels have been published previously^9^.

CoMix was first launched in March 2020 in the UK, Belgium, and the Netherlands^10,11^. In the UK, two panels of respondents are asked to respond once every two weeks, in alternating weeks. Initially, each panel consisted of about 1500 participants, increasing to about 2500 participants each week from August 2020. We recruited new participants on a rolling basis as existing participants dropped out of the study. Participants were included for a maximum of 7-10 survey rounds. Further details of CoMix in the UK have been published elsewhere^10,12^. In the other CoMix countries the sample size were smaller. To the original three CoMix countries, initially 7 countries (Austria, Denmark, France, Italy, Poland, Portugal, and Spain) between December 2020 and April 2021, then 6 countries between February 2021 and October 2021 (Finland, Greece, Lithuania, Switzerland, and Slovenia), and lastly 4 countries between May 2021 and October 2021 (Hungary, Slovakia, Estonia, and Croatia) were added^7^. In each of these countries, the initial panel consisted of at least 1500 participants, who were invited to 7 survey rounds. The original CoMix survey was translated into the local languages (available on https://github.com/wongkerry/comix_preg/questionnaire).

### Study participants

In this analysis, we included pregnant women (self-identified) aged 18-49 years, and non-pregnant women and men of the same age who reported to have no risk factors for serious symptoms if they contracted COVID-19. We used self-reported pregnancy status to identify pregnant women. Women who had reported not being pregnant for all surveys they completed were considered non-pregnant.

### Data

We combined data on social contacts, risk perception, status, mitigation, and COVID-19 vaccination from the participants of the CoMix survey and information on non-pharmaceutical interventions (NPI) in the study countries from the Oxford Coronavirus Government Response Tracer (OxCGRT) project^13^, which provides a systematic way to track government responses to COVID-19 across countries.

#### Reporting of contacts (CoMiX) and isolation or quarantine due to COVID-19

Participants reported social contacts that occurred on the day prior to the survey. The details of reporting have previously been documented^10^. Briefly, participants reported contacts by listing each individual or as a total number of contacts made by setting. A direct contact is defined as anyone who met the participant in person with whom at least a few words were exchanged in close proximity, or physical contact was made. We grouped reported contacts by the settings in which they were made – at home and outside of the home setting; we further distinguished contacts made outside of the home setting as contacts made at work, contacts made in social settings and contacts made elsewhere. Social settings were taken as someone else’s house, a place of worship, at a shop for non-essential items, at a place of entertainment such as a restaurant, bar, cinema, at a place for sports, and other outside locations such as in a park or in the countryside. We also asked if participants had been in “isolation or quarantine due to coronavirus (Covid-19)?” in the last seven days. Isolation or quarantine could include staying at home after potential exposure to an infected case, on return from a trip abroad, or separation from people who are not infected, either at home or in a facility.

#### Risk perception, status, mitigation and COVID-19 vaccination (CoMiX)

In addition to social contacts, participants were asked to respond to statements regarding their perception of risk. Participants were asked to respond to the statements:

i. “I am likely to catch coronavirus”,
ii. “I am worried that I might spread coronavirus to someone who is vulnerable”, and
iii. “Coronavirus would be a serious illness for me”.

The responses were captured with the likert scale of “Strongly Agree”, “Tend to Agree”, “Neutral”, “Tend to Disagree”, and “Strongly Disagree”. Participants were also asked whether they wore a face covering.

Since December 2020, we added questions on vaccination against COVID-19. Participants were asked “Have you been vaccinated against the virus that causes Coronavirus (COVID-19)?”, or “Have you had any new doses of the vaccination since you last completed the survey?”. From the day of survey response, participants who responded they had received the vaccination in either of these two questions were considered vaccinated (either partially vaccinated or fully vaccinated).

#### NPIs (OxCGRT)

We extracted data on NPIs from the OxCGRT project^13^ as measured by an overall stringency index (SI) in each of the study counties. SI is calculated using eight containment and closure policies (school closing, workplace closing, cancelling of public events, restrictions on gathering sizes, closing public transport, stay at home requirements, restrictions on internal movement and international travel controls), plus an indicator recording public information campaigns. SI takes values from 0 (least strident restrictions) to 100 and provides a systematic way to quantify the strictness of “lockdown style” policies aimed primarily at restricting behaviours. In April 2020 in the UK, for instance, all four nations were in the lockdown with education establishments, non-essential retail, alongside the leisure and hospitality sectors closed. People were required to only leave their house for essential shopping or medical needs, or in order to undertake one form of exercise per day. SI at the time was 80. In July 2021, on the other hand, SI was approximately 30 when only few restrictions were in place (Supplementary Material I).

### Statistical analysis

#### Descriptive

R version 4.0.5 was used for all analyses and the code and data are available on https://github.com/wongkerry/comix_preg.

We presented summary statistics of age, household size, employment status, and occupation (available only in some countries) of the study sample by pregnancy status.

We calculated the predicted mean contacts in different settings (all settings, home, outside of home, work, and other social settings) using a generalized additive mixed model (GAMM)^14,15^. We assumed reported contacts followed a negative binomial distribution, modelled using a log link function, with a random effect for participants by pregnancy status and gender. The predictions were adjusted for employment status (full-time, part-time, and self-employed) and year-month, and weighted by weekday.

The proportion and associated confidence intervals of participants in isolation or quarantine, agreed/strongly agreed to statements related to perception of risk, the use of face-covering, and vaccination coverage over time with 1000 samples using clustered bootstrapping^13^. Each participant was sampled with replacement and then all observations for selected participants were included in bootstrapped samples to account for dependency from repeated observations of the same participants. This calculation was based on a moving window over two-weeks, overlapping intervals to increase the sample size per estimate and to include all participants from simultaneously running panels. We assess vaccination coverage in all countries except for the UK together due to small sample size in each of the other individual countries. The status of vaccination policy for pregnant women between March and September 2021 in each country is given in Supplementary Material III^16^.

## Results

### Participant characteristics

Overall, we recorded surveys completed by 30,901 participants aged 18-49 years between March 2020 to September 2021, 1,041 (3.4%) of whom were pregnant women (Table 1). The mean age was approximately 33.8 years in all groups. Over half (54.8%) of all participants reported to be in full-time employment, with a greater proportion of men reported being employed fulltime (64.1%). The mean household size was 3 people.

**Table 1.**
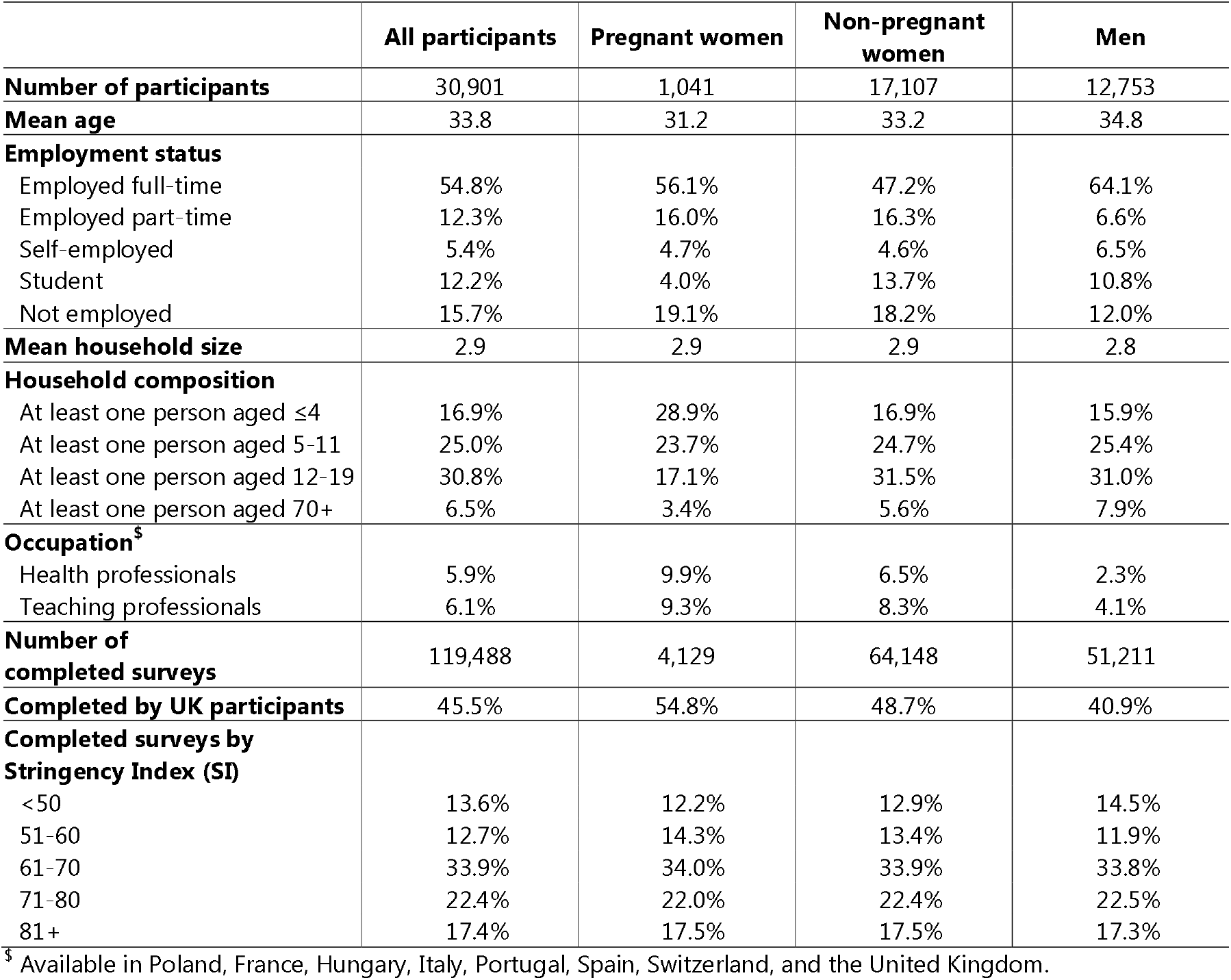
Characteristics of study participants and distribution of completed surveys by country and stringency index of NPIs

Over the entire study period, the participants completed 119,488 surveys, 4,129 of which by pregnant women. Nearly half (45.5%) of the surveys were completed by participants in the UK, with each of the other 18 countries contributing between 1.0% to 5.2% (Supplementary Material II). Prior to late-December in 2020, the study sample was based in the UK, Belgium, and the Netherlands (Figure 1). The study sample included more surveys completed by participants from other European countries since late-December 2020 – e.g., Italy, France, Spain, Denmark, Poland, and Austria.

**Figure 1.**
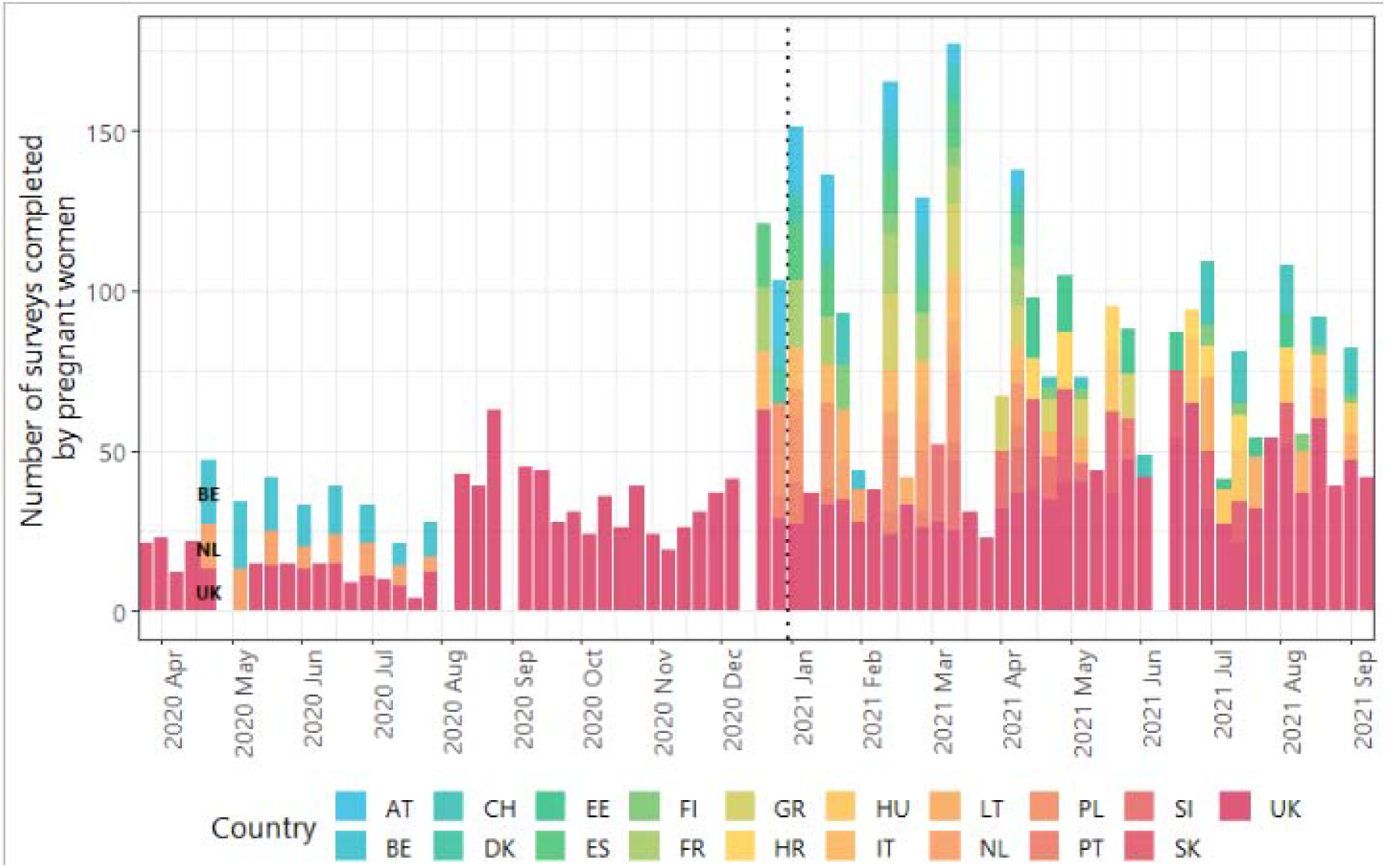
Number of surveys completed by pregnant women over time by country between 23 March 2020 and 12 September 2021 Not all countries are labelled. The dashed line represents 1^st^ January 2021.

Nearly half (39.8%) of the surveys were completed when NPIs were stringent (Stringency Index (SI) of 71+), with 13.6% completed when most restrictions were relaxed, and SI was low.

### Mean contacts

The mean number of daily contact reported over the entire study period was slightly lower in pregnant women (3.6, 95%CI=3.5-3.7) than in non-pregnant women aged 18-49 (4.0, 95%CI=3.9-4.0), and men of same age (3.7, 95%CI=3.7-3.8) (Table 2). Contacts at home were approximately 1.5 in all groups. Contacts made at work were lower among pregnant women (0.9, 95%CI=0.8-1.1) than in non-pregnant individuals (1.3-1.4). Contacts made at social settings and outside of the participants’ household were higher among pregnant women (0.7, 95%CI=0.6-0.7) than in non-pregnant people (0.5-0.6).

**Table 2.** Bootstrap crude mean contacts with 95% confidence interval of bootstrapping

|  | Pregnant women | Non-pregnant women aged 18-49 | Men aged 18-49 |
| --- | --- | --- | --- |
| **All settings** | 3.6 (3.5-3.7) | 4.0 (3.9-4.0) | 3.7 (3.7-3.8) |
| **Contacts at home** | 1.6 (1.5-1.6) | 1.6 (1.6-1.6) | 1.5 (1.4-1.5) |
| **Contacts not at home** | 2.1 (1.9-2.2) | 2.3 (2.3-2.4) | 2.3 (2.2-2.3) |
| At work | 0.9 (0.8-1.1) | 1.3 (1.3-1.4) | 1.4 (1.3-1.4) |
| At social settings | 0.7 (0.6-0.7) | 0.6 (0.6-0.6) | 0.5 (0.5-0.5) |

#### Contacts by setting and NPIs

Contacts varied by settings as well as the strictness of the NPIs that were put in place. Home contacts remained consistent at 1.0-2.5 for pregnant women, non-pregnant women and men across all levels of NPI strigency (Figure 2), contacts not at home declined in all groups. Pregnant women generally made the least contacts outside of the home setting at all levels of NPIs, driven primarily by fewer contacts made at work – in most cases <2 and at high SI <1 work contacts vs. 2-3 among non-pregnant people. In social settings, however, pregnant women reported more contacts than non-pregnant people at all levels of restrictions.

**Figure 2.**
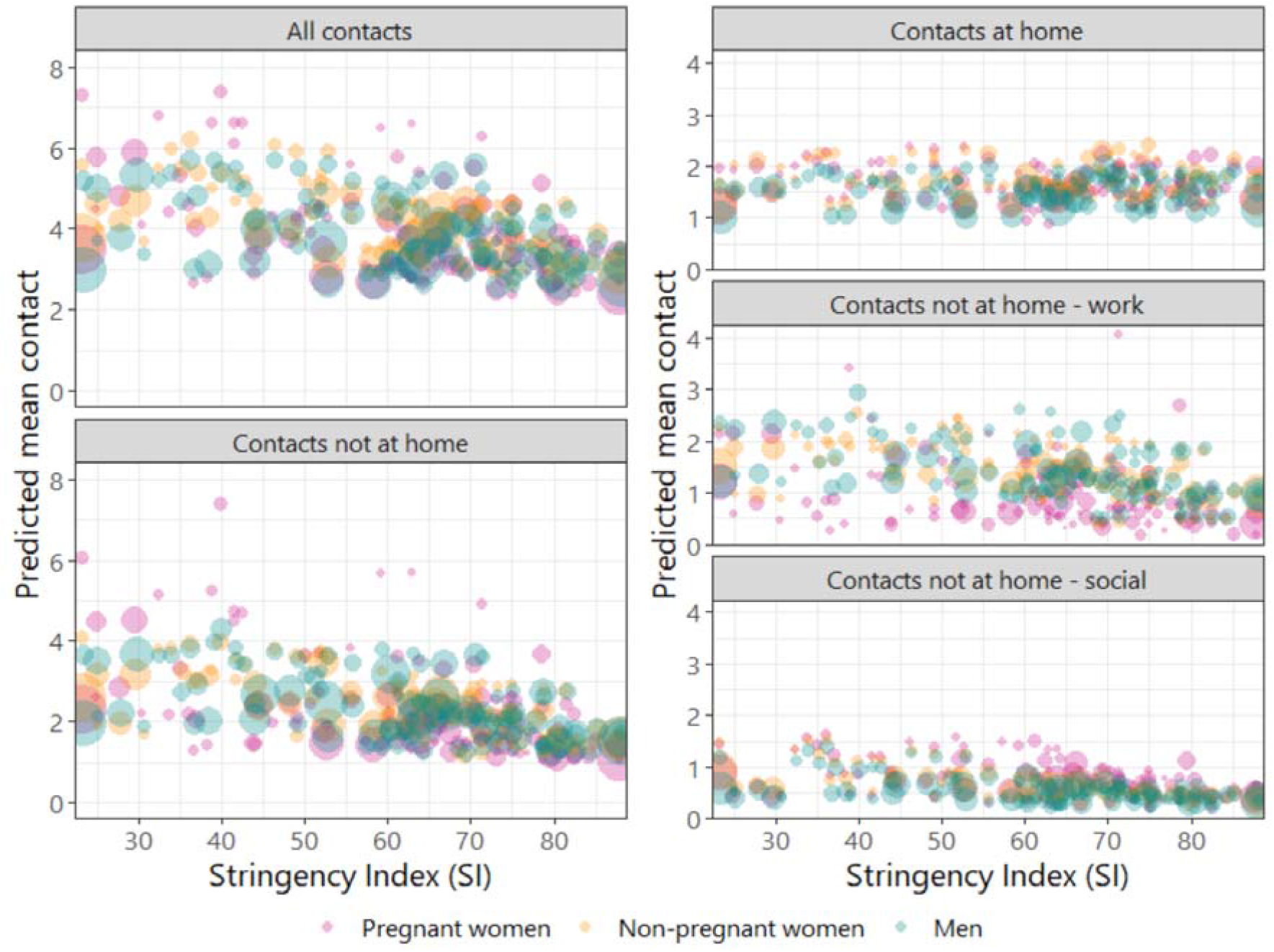
Predicted mean contact in different settings and by different levels of the Stringency Index Predicted mean contact is adjusted for employment status and month. Every point represents one observation in a country. Size of the bubbles is scaled to the number of completed surveys by pregnancy status and sex.

Further to making generally fewer contacts, a consistently greater proportion of pregnant women reported being in isolation or quarantine due to COVID-19 – approximately 20%-40% pregnant women versus <20% in non-pregnant women and men prior to May 2020, and 15% versus 5% since May 2020 (Figure 3).

**Figure 3.**
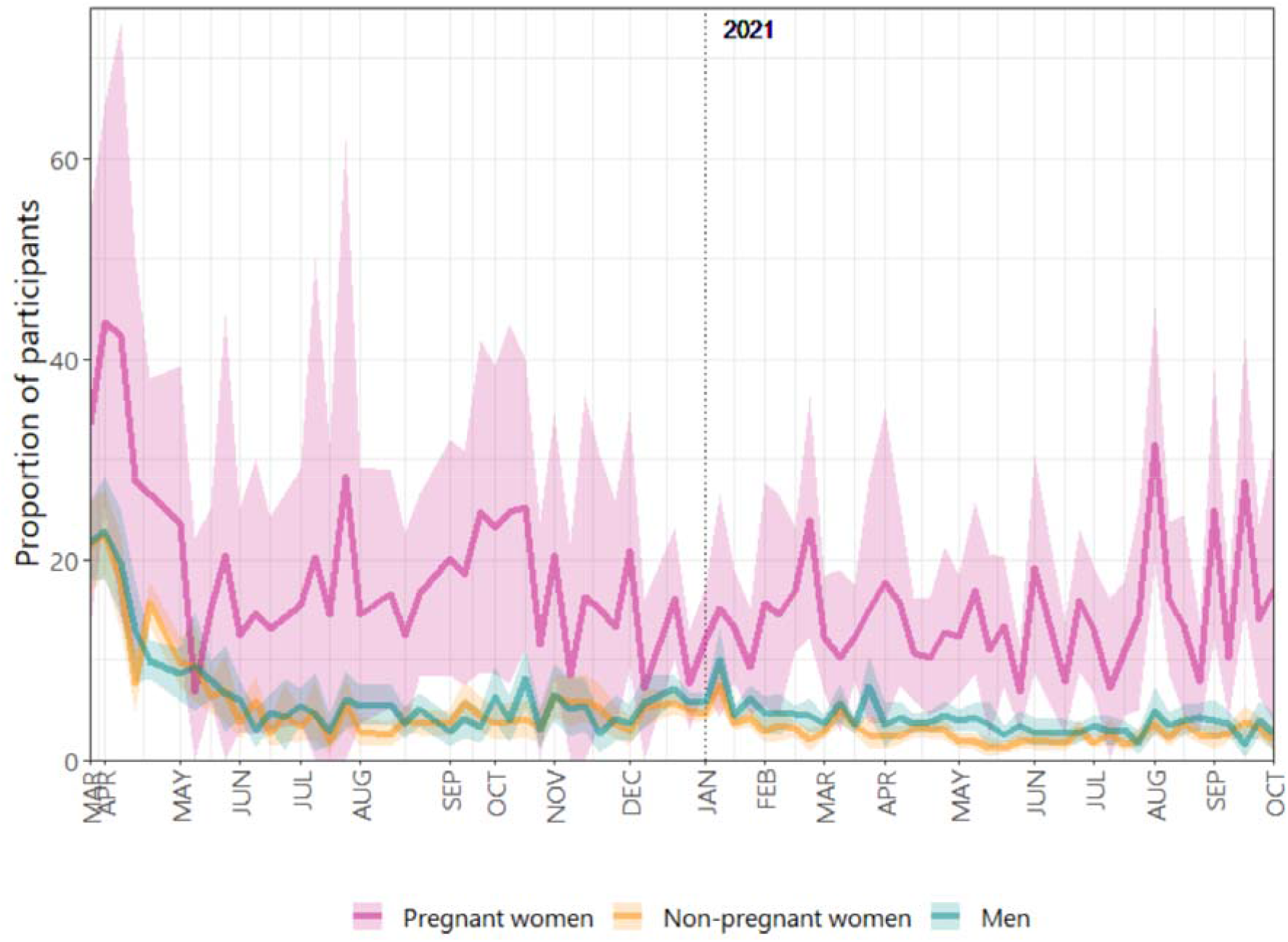
Proportion of participants who report being in isolation or quarantine due to COVID-19 with 95% bootstrap-based confidence interval.

### Risk Perception and use of face-covering

Approximately 45-50% of pregnant women answered “Agree” and “Strongly agree” to a statement indicating that coronavirus would be a serious illness for them throughout the entire study period (Figure 4a). On the other hand, the proportion of non-pregnant people who agreed and strongly agreed with the statement dropped from approximately 35% and maintained at approximately 25% since August 2020.

**Figure 4.**
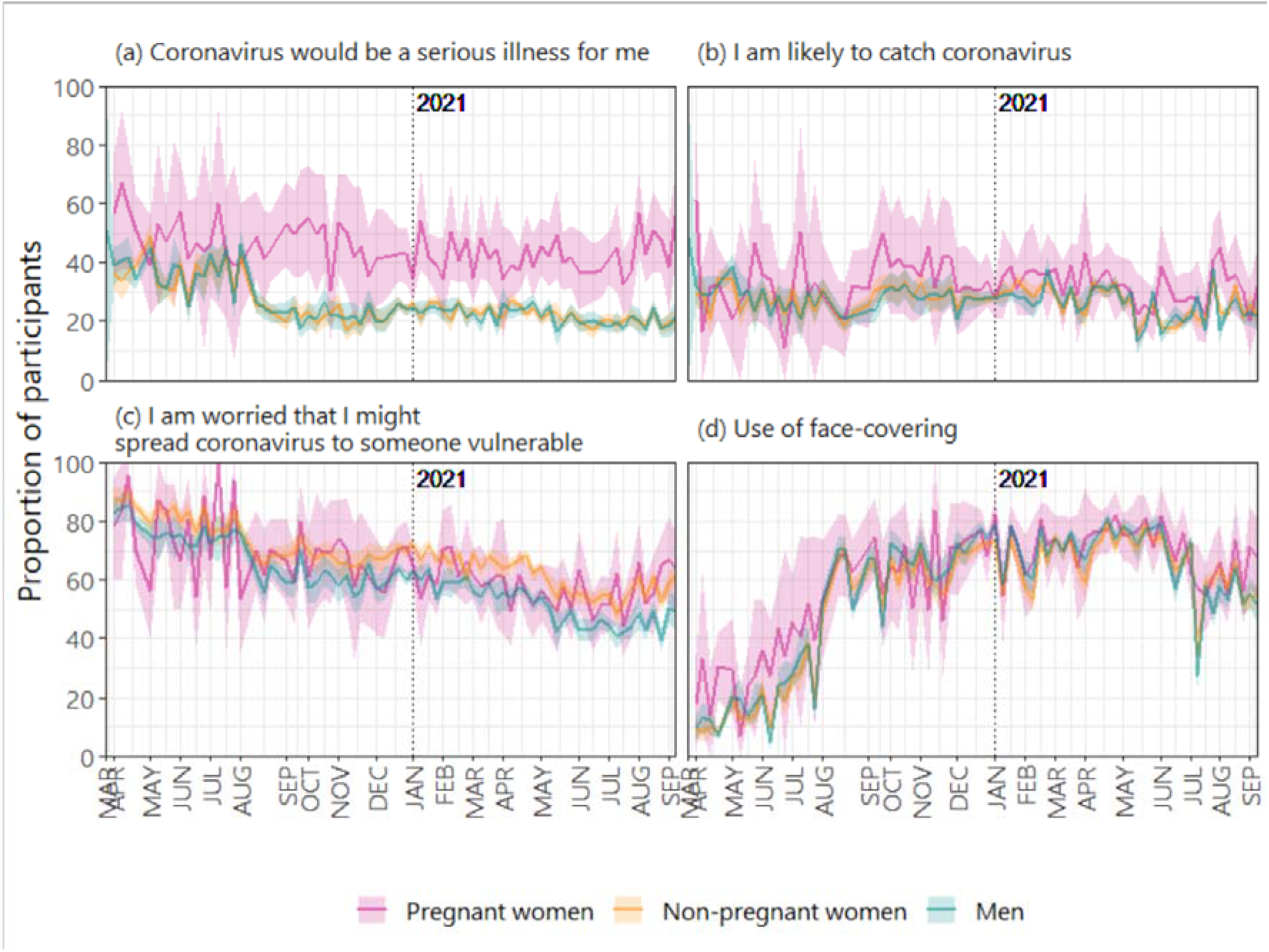
The proportion of participants who answered; with bootstrap-based 95% confidence intervals.

The proportion who answered “Agree” and “Strongly agree” to the statement “I am likely to catch coronavirus” remained consistent at roughly 25% over time and across both pregnant and non-pregnant individuals (Figure 4b). The proportion of participants who were worried that they might spread coronavirus to someone vulnerable declined over time in all groups (Figure 4c). There was some evidence to suggest that slightly more non-pregnant women were more worried that they might spread coronavirus to someone vulnerable (as compared to men).

The use of face covering was 15-20% points higher among pregnant women (30-50%) compared to non-pregnant people between April and August 2020 (Figure 4d). There was a sharp increase to roughly 70% use of face-covering reported by all groups in August 2020. Usage of face-covering then remained consistent in all groups until a decline to 60% since July 2021.

### Vaccination against COVID-19

We plotted the monthly vaccination coverage among pregnant women against that of non-pregnant women (Figure 5). Between January and March 2020, vaccine coverage was <10% in both groups of women in 18 European countries (although only 7 countries contributed data to this period). Since May 2021, vaccination coverage in non-pregnant women began to increase whilst coverage in pregnant women remained broadly static. In June, for instance 55% of non-pregnant women in European countries were partially or fully vaccinated. This contrasted with 25% of pregnant women being partially or fully vaccinated. A similar pattern was also observed among UK participants (Figure 5).

**Figure 5.**
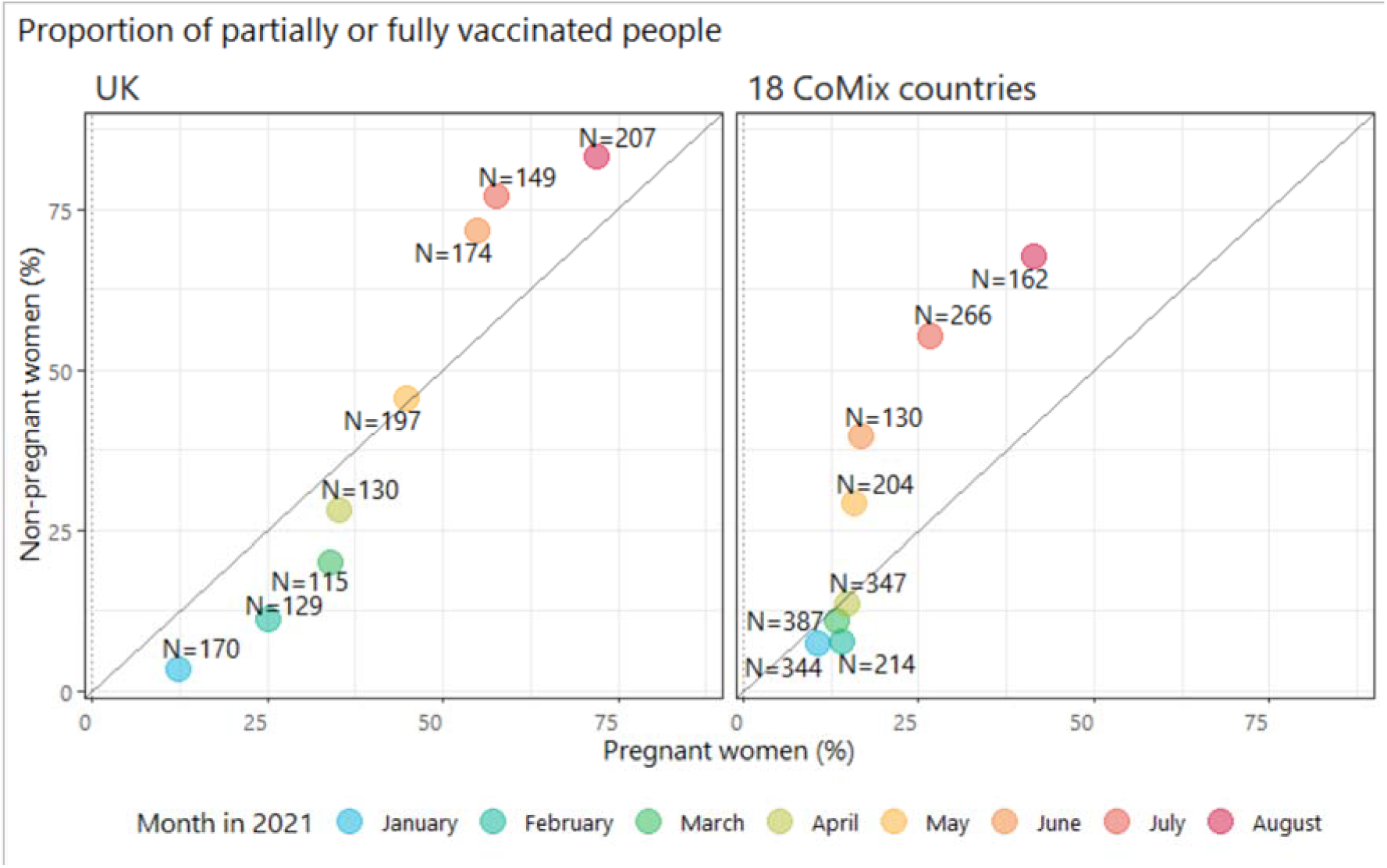
Monthly proportion of pregnant and non-pregnant women who report having been fully or partially vaccinated against COVID-19 in 18 European countries and in the UK Note: The numbers on the graph indicate the numbers of surveys completed by pregnant women in that month.

## Discussion

Connections with and care from people in one’s support network help regulate emotions, cope with stress, and remain resilient, and may be viewed particularly important for women entering parenthood or extending their family. Since the COVID-19 pandemic in spring 2020, however, pregnant women have had to navigate additional challenges inherent to the pregnancy period with reductions of social support in the context of sudden, severe, and stressful events. This is further complicated by worse clinical outcomes for both mothers and babies who have COVID-19, and the lack of a consensus on the COVID-19 vaccine by government and health authorities around the world. Pregnant women have reported greater COVID-19 related fear and concerns since the start of the pandemic^4^, and this study confirms their heightened worry about getting severely ill from COVID-19 compared to their non-pregnant counterparts, pointing to even greater need for psychosocial care during the vulnerable period of childbearing^17^.

Prenatal stress may pave the way to postnatal symptoms of depression and anxiety for the mother, and worse behavioural, emotional, cognitive, and physical outcomes for the children^18^. Social and physical distancing likely threaten to aggravate the feelings of concern and isolation that will produce these negative health consequences^19^. In this study, pregnant women maintained a similar level of contacts in “non-essential” social settings – such as other people’s home, a place of entertainment or sports – regardless of NPIs, suggesting a minimum level of social contacts that they may consider as vital. Additional attention by healthcare workers and dedicated programs can help care for maternal mental health, especially in settings of high rates of COVID-19 infection and strict NPIs. While the current study focused on the pregnancy period, continued support in the postpartum period is also essential for ongoing maternal, child and family health.

Pregnant and reproductive-aged women account for a large proportion of the population with particular concerns regarding vaccination against COVID-19. Recent studies conducted in UK, Ireland, and the United States revealed greater vaccine hesitancy in pregnant women compared to non-pregnant women of the same age^20,21^. This pattern is reflected in our findings (apart from the first few months of the vaccination programme when healthcare workers were among the prioritized groups to receive the vaccine). Concerns about how rapidly the vaccines were developed, safety, side effects, and possibility of harm to the foetus were frequently raised^22^.

We combined data on vaccine coverage in 18 different European countries – due to low number of participants from this vulnerable group in each individual country – bearing in mind the different COVID-19 maternal immunization policies across countries, and over time in a country. The finding on lower vaccine coverage in pregnant women than in non-pregnant women is likely due to a mixture of policies barring access to vaccines based on pregnancy status and low uptake by pregnant women where the vaccine is offered. So far, evolving recommendations, and misinformation have led to some understandable hesitancy among pregnant women. As more evidence about the safety and effectiveness of COVID-19 vaccines in pregnancy becomes available and provides increasing reassurance that the benefits of vaccination (especially in settings with high rates of COVID-19 infection) is in favour of vaccination for pregnant women^23^, advice from those responsible for maternal and neonatal health in many countries is that pregnant women should consider COVID-19 vaccination^24^. Priority should be given to informing pregnant women and healthcare professionals on vaccine safety and effectiveness, alongside strategies to address vaccine hesitancy, including post-vaccination surveillance and long-term mother and infant follow up. The complex nature of vaccine acceptance is evidenced by further divides between different socio-demographic subgroups^17^. Better understanding of these concerns is important to devising targeted approaches to all subgroups.

The findings presented in this study should be interpreted with several limitations in mind. First, more than half of our data came from the UK which may have obscured patterns from other countries. Relatedly, for the analysis of COVID-19 vaccination in countries other than the UK, we grouped 18 countries together due to data availability. This approach neglected variations in vaccination schedules and the evolution of vaccine policies on pregnancy in individual countries. Both risk perceptions to COVID-19 and vaccine hesitancy also likely differ across countries and change over time. Future research is highly warranted as more data become available. Second, we did not collect data on gravidity and gestational age from pregnant women, which likely have important relationships with perceived need of social support and vaccine acceptance. Future studies should explore factors associated with subgroups of pregnant women requiring particular attention. Third, pregnant and non-pregnant individuals may have interpreted the questions on isolation and quarantine due to COVID-19 differently. Terminologies such as “isolation”, “quarantine” and “social distancing” are distinctively different yet somewhat similar notions that might be misinterpreted, especially for high-risk people who have been given stricter guidelines to maintain social distancing as a form of preventive measure. Our finding on isolation and quarantine might have over-estimated the proportion of pregnant women who were truly in isolation or quarantine. Fourth, the study was conducted online using a quota-based sample of individuals who had agreed to participate in a marketing survey. The recruitment method may not lead to a representative sample of pregnant women, and is particularly prone to bias towards people with access to the internet and who may be reached by online advertising campaigns. The other limitations of our online survey have been documented elsewhere^7,10^.

## Conclusion

The COVID-19 pandemic quickly transformed the global social landscape as widespread NPIs reshaped social behaviour, especially for the most vulnerable. The vigilance with social behavioural guidelines given to pregnant women has limited their access to seek in-person support from others during pregnancy, typically a time of heightened social need. Furthermore, the confusion surrounding pregnant women has likely interfered with decision making about the COVID-19 vaccination. Recognition of maternal psychosocial wellbeing and, strengthening the delivery of clear and correct information may offer reassurance to women whilst pregnant during the COVID-19 pandemic.

## Data Availability

All data produced in the present work are contained in the manuscript.

## Declaration section

### Ethics approval and consent to participate

Participation in this opt-in study was voluntary, and all analyses were carried out on anonymised data. The corresponding author confirm that all experimental protocols were approved by the ethics committee of the London School of Hygiene and Tropical Medicine Reference number 21795. The corresponding author confirm that all methods were carried out in accordance with relevant guidelines and regulations. The correspond author confirm that informed consent was obtained from all subjects and/or their legal guardians.

### Consent for publication

Not applicable

### Availability of data and materials

The datasets generated and analysed the current study is available from http://www.socialcontactdata.org/data/.

### Funding

The following funding sources are acknowledged as providing funding for the named authors.

HPRU in Modelling & Health Economics (NIHR200908: KLMW);

European Union Horizon 2020 research and innovation programme – (EpiPose 101003688: AG, WJE) Wellcome Trust (213589/Z/18/Z: ESP)

European Research Council (ERC) under the European Union’s Horizon 2020 research and innovation programme (TransMID 682540: CF, PB, NH)

This research was partly funded by the Global Challenges Research Fund (GCRF) project RECAP managed through RCUK and ESRC (ES/P010873/1: CIJ)

NIHR (PR-OD-1017-20002: WJE)

UK MRC (MC_PC_19065 - Covid 19: Understanding the dynamics and drivers of the COVID-19 epidemic using real-time outbreak analytics: WJE)

The funders had no role in study design, analysis, decision to publish or preparation of the manuscript.

### Authors’ contributions

KLMW and WJE conceived of and planned the analysis; KLMW performed the main analysis based on methods previously developed by AG, CIJ, and WJE; CIJ and WJE designed the CoMix contact survey, CIJ, AG, and KLMW cleaned and managed the contact survey data; All authors wrote and reviewed the manuscript. The CoMix Europe Working Group provided discussion and comments.

## Acknowledgement

The following authors were part of the CoMix Europe Working Group. Each contributed to consultation for policy input, interpretation of data and findings, and supported the drafting of the manuscript, and approved the work for publication:

- Daniela Paolotti, Michele Tizzani and Ciro Cattuto (Data Science for Social Impact, ISI Foundation)
- André Karch, and Veronika Jäger (Institute of Epidemiology and Social Medicine, University of Münster)
- Andrea Schmidt, Gerald Gredinger, and Sophie Stumpfl (Health Economics and Health System
- Analysis, Austrian National Public Health Institute)
- Joaquin Baruch, and Tanya Melillo (Infectious Disease Prevention & Control Unit, Ministry for Health, Malta)
- Henrieta Hudeckova, Jana Zibolenova, and Zuzana Chladna (Department of Public Health, Comenius University Bratislava, Jessenius Faculty of Medicine in Martin)
- Magdalena Rosinska, and Marta Niedzwiedzka-Stadnik (Department of Infectious Disease Epidemiology and Surveillance, National Institute of Public Health NIH - National Research Institute, Poland)
- Krista Fischer, Sigrid Vorobjov, and Hanna Sõnajalg (Department of drug and infectious
- diseases epidemiology, National Institute for Health Development, Estonia)
- Christian Althaus, Nicola Low, and Martina Reichmuth (Institute of Social and Preventive Medicine, University of Bern)
- Kari Auranen, and Markku Nurhonen (Department of Health Security and Dept. of Public Health and Welfare, National Institute for Health and Welfare, Finland)
- Goranka Petrović, and Zvjezdana Lovric Makaric (Division for Epidemiology of Communicable Diseases, Croatian Institute of Public Health)
- Sónia Namorado, Constantino Caetano, and Ana João Santos (Department of Epidemiology, National Institute of Health Dr Ricardo Jorge, Portugal)
- Gergely Röst, Beatrix Oroszi, and Márton Karsai (Bolyai Institute, University of Szeged)
- Mario Fafangel, Petra Klepac, and Natalija Kranjec (Communicable Diseases Centre, National Institute of Public Health, Slovenia)
- Cristina Vilaplana, and Jordi Casabona (Experimental Tuberculosis Unit, Institut d’Investigació en Ciències de la Salut Germans Trias i Pujol, Spain)

We acknowledge support from the European Centre for Disease Prevention and Control (ECDC) in setting up the collaborations between the Epipose consortium, and universities and public health institutions in all other countries. We gratefully acknowledge the tremendous efforts put in all the steps by the EpiPose consortium, its collaborators and Ipsos.

We would also like to thank the team at Ipsos who have been excellent in running the survey, collecting the data, and allowing for this study to, happen at a rapid speed. We acknowledge the exceptional project management support given by Sarah Vercruysse and Bieke Vanhoutte.

