## Supplementary material for "Pregnancy during COVID-19: social contact patterns and vaccine coverage of pregnant women from CoMix in 19 European countries": Supp I-III (from another version of this preprint: version 1, 2022-06-02)

**Supplementary Material I** Oxford COVID-19 Government Response Tracker Stringency Index in 21 study countries

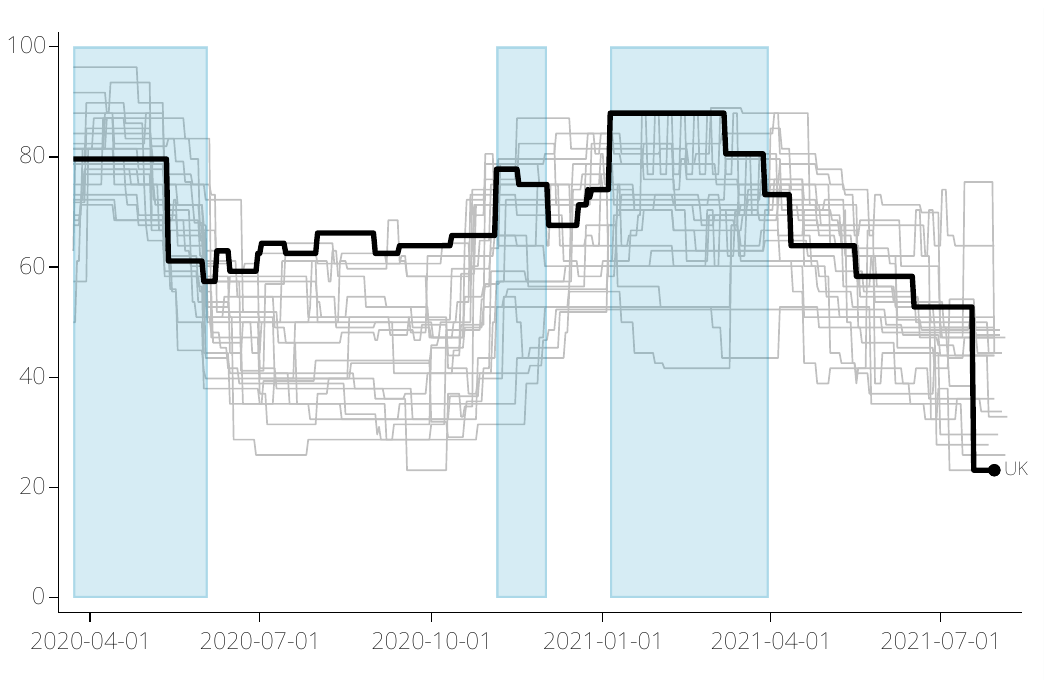

**Supplementary Material II** Number of participants and number of completed surveys by country

|  | **Number of participants aged 18-49** | | | | **Number of surveys completed by participants aged 18-49** | | | |
| --- | --- | --- | --- | --- | --- | --- | --- | --- |
|  | **All participants** | **Pregnant**  **women** | **Non-pregnant women** | **Men** | **All participants** | **Pregnant**  **women** | **Non-pregnant women** | **Men** |
| **By country** |  |  |  |  |  |  |  |  |
| DK (Denmark) | 622 | 12 | 328 | 282 | 2330 | 44 | 1200 | 1086 |
| PT (Portugal) | 889 | 14 | 492 | 383 | 3168 | 50 | 1733 | 1385 |
| IT (Italy) | 670 | 18 | 291 | 361 | 3026 | 88 | 1310 | 1628 |
| ES (Spain) | 682 | 20 | 337 | 325 | 3015 | 99 | 1500 | 1416 |
| FI (Finland) | 1083 | 20 | 558 | 505 | 4102 | 60 | 1968 | 2074 |
| HR (Croatia) | 924 | 22 | 505 | 397 | 3229 | 81 | 1687 | 1461 |
| AT (Austria) | 878 | 23 | 477 | 378 | 3223 | 85 | 1708 | 1430 |
| EE (Estonia) | 755 | 26 | 366 | 363 | 2744 | 83 | 1219 | 1442 |
| FR (France) | 720 | 26 | 290 | 404 | 2774 | 114 | 1088 | 1572 |
| HU (Hungary) | 954 | 28 | 500 | 426 | 3049 | 97 | 1539 | 1413 |
| NL (Netherland) | 1211 | 29 | 608 | 574 | 5212 | 130 | 2564 | 2518 |
| BE (Belgium) | 1341 | 31 | 743 | 567 | 5189 | 150 | 2752 | 2287 |
| GR (Greece) | 1284 | 33 | 691 | 560 | 3756 | 111 | 1980 | 1665 |
| PL (Poland) | 824 | 33 | 441 | 350 | 3182 | 153 | 1756 | 1273 |
| SI (Slovenia) | 1108 | 34 | 645 | 429 | 3659 | 110 | 2181 | 1368 |
| SK (Slovakia) | 837 | 35 | 413 | 389 | 3219 | 147 | 1490 | 1582 |
| LT (Lithuania) | 1312 | 42 | 792 | 478 | 5032 | 133 | 2712 | 2187 |
| CH (Switzerland) | 2008 | 49 | 989 | 970 | 5199 | 132 | 2575 | 2492 |
| UK (United Kingdom) | 12799 | 546 | 7641 | 4612 | 54380 | 2262 | 31186 | 20932 |
| **Total** | **30901** | **1041** | **17107** | **12753** | **119488** | **4129** | **64148** | **51211** |

(The rows are sorted by the number of pregnant participants aged 18-49)

**Supplementary Material III** Covid-19 vaccine policies on pregnancy in 18 CoMix countries between March and September 2021. For each country, we extracted one data point per month from https://www.comitglobal.org/. Data is available from March 2021.
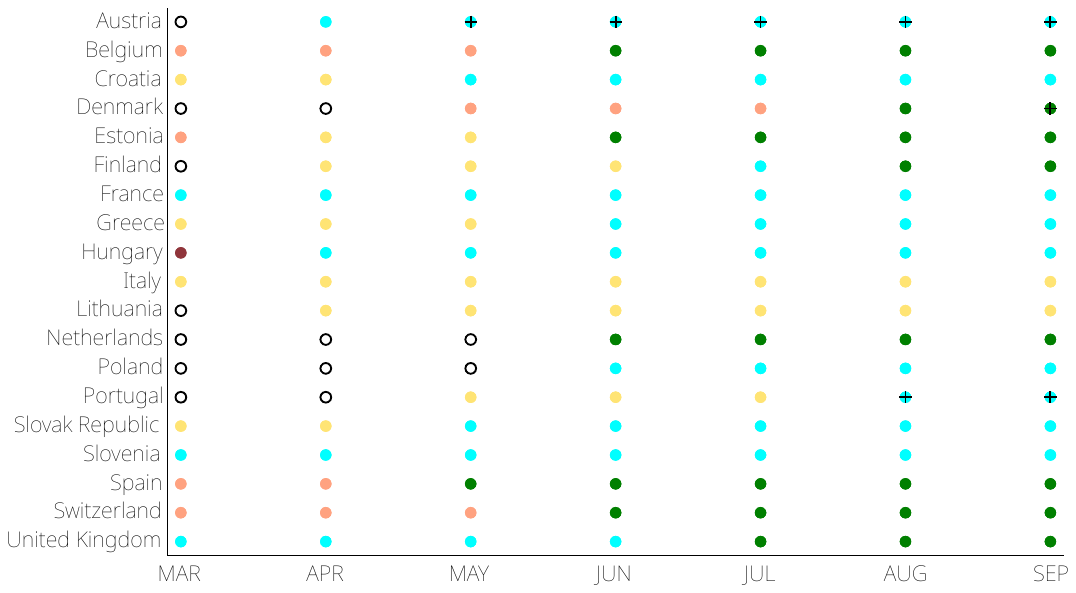

| ● | **Recommended for some or all**  An explicit recommendation that some or all pregnant people should receive vaccine. |
| --- | --- |
| ● | **Permitted**  All pregnant people can receive, may receive, or can choose to receive vaccine. |
| ● | **Permitted with qualifications**  Only certain groups of pregnant people, e.g., pregnant health workers, pregnant people with underlying conditions, can, may, or can choose to receive vaccine. |
| ● | **Not recommended but with exceptions**  A statement stating pregnant people should not receive vaccine, with certain exceptions. |
| ● | **Not recommended**  People who are pregnant should not receive the vaccine or vaccine is contraindicated. |
| ● | **No policy found**  No policy/position found regarding vaccinating pregnant people can be found, or where no position was clearly established. |
| ○ | **No data** |
| + | **Advised for pregnant women who are in their 2^nd^ and 3^rd^ trimester** |
